# Detection of *P. malariae* using a new rapid isothermal amplification lateral flow assay

**DOI:** 10.1101/2023.02.26.23286371

**Authors:** Ashenafi Assefa, Kevin K. Wamae, Chris M. Hennelly, Billy Ngasala, Meredith Muller, Albert Kalonji, Fernandine Phanzu, Clark H. Cunningham, Jessica T. Lin, Jonathan B. Parr

**Author notes:** **Correspondence:** Ashenafi Assefa, University of North Carolina at Chapel hill, Chapel Hill, North Carolina, USA. or.

## Abstract

*P. malariae* is found worldwide and causes chronic parasitism in its human hosts. We developed a *P. malariae (Pm)* diagnostic assay that uses rapid, isothermal recombinase polymerase amplification (RPA) and lateral-flow-strip detection. Using 18S rRNA plasmid DNA, the assay demonstrates a detection limit of 10 copies /µL (∼1.7 genome equivalents) and 100% analytical specificity. Testing in field samples showed 95% clinical sensitivity and 88% specificity compared to qPCR. Total assay time was 35 minutes. Combined with simplified DNA extraction methods, the assay has potential for future field-deployable point-of-care use to detect a parasite species that remains largely undiagnosed.

## INTRODUCTION

Intensive malaria control efforts have yielded progress toward malaria elimination in multiple endemic countries [1]. In some settings, as the burden of *Plasmodium falciparum* (Pf) and *P. vivax* (Pv) malaria declines, less common species such as P. malariae (Pm) and P. ovale (Po) have increased in prevalence as well as public health relevance [2]. Malaria elimination requires rapid detection and treatment of *all Plasmodium* species. However, existing rapid methods are not species-specific and have poor sensitivity for these less common species [3]. Though *P. malariae* is known to cause chronic parasitemias, sometimes lasting years, and causing chronic anemia and splenomegaly [4], its treatment is straightforward. Thus, its detection and treatment can reduce chronic carriage and contribute to malaria elimination efforts. This study reports a new Pm-specific diagnostic assay that uses rapid, isothermal recombinase polymerase amplification (RPA) and lateral flow strip detection and has potential for further development into a point-of-care tool.

## MATERIALS AND METHODS

### Primer and Probe Design

Primers and probes were designed according to manufacturer-suggested best practices [TwistDX (TwistAmpTM nfo), Cambridge, United Kingdom]. Primers were designed to meet the following parameters: 30-36 nucleotide (nt) length, 40-50% GC content, 50-100°C melting temperature, <5nt mononucleotide repeat length, and 80-500nt amplicon length. Publicly available Pm 18S ribosomal RNA gene sequences from PlasmoDB and NCBI databases were used to design primers using Primer3Plus v3.2. Primers from published literature were also modified and used to guide selection of target regions [5-7]. Probes were designed within the target sequence, with 46-52nt length, 20-80% GC content, and 57–80°C melting temperature.

### RPA assay protocol

RPA reactions were performed using a reverse primer biotinylated on the 5’ end and an unlabeled forward primer; a 5’-FAM-labelled probe with an abasic residue and 3’ blocker modification. The blocker prevents extension until cleavage of the abasic site by Endonuclease IV (nfo) enzyme. The biotinylated primer and FAM labelled probe form a duplex of double-stranded RPA amplicons that are detectable by sandwich assay, involving a lateral flow strip test band containing anti-FAM antibodies. Direct visualization by naked eye is possible using streptavidin-gold conjugates that bind biotinylated double stranded amplicon captured by anti-FAM antibodies on the lateral flow strip (**Figure 1**).

**Figure 1.**
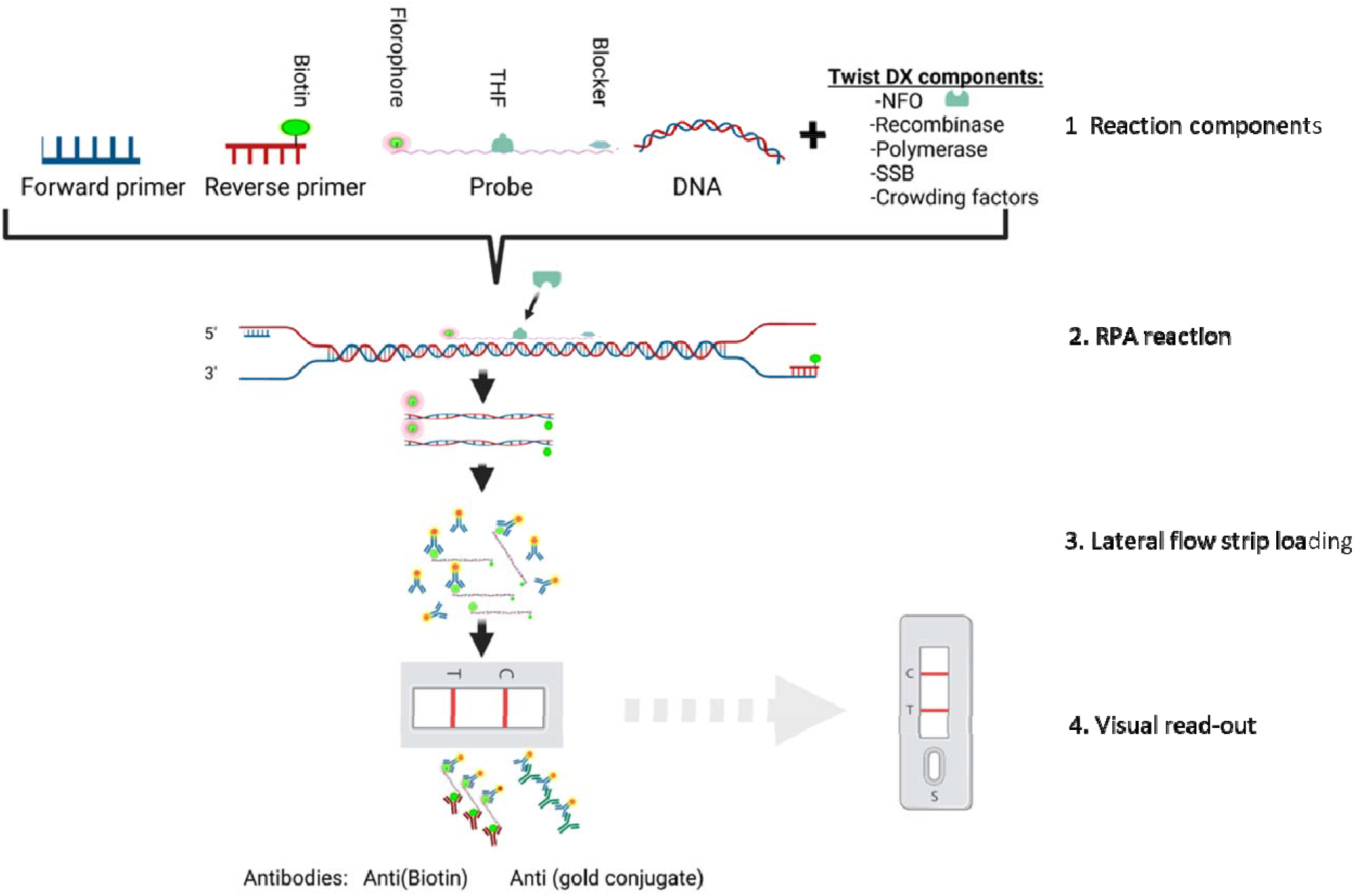
*P. malariae* lateral-flow based RPA assay schematic. 1) Reaction components include unlabeled forward and 5’-biotinylated reverse primers, 5’-FAM labeled probe with an abasic residue and 3’ blocker, endonuclease IV enzyme (nfo), and template DNA. 2) RPA reaction proceeds via primer-recombinase-SSB complex de-looping, nfo cleavage, polymerase extension that results in FAM- and biotin-labeled double-stranded amplicons. Exponential amplification occurs during 20-minute isothermal incubation at 39°C. 3) Lateral flow strips with a band containing anti-FAM antibodies are loaded with RPA product diluted in buffer containing streptavidin-conjugated gold nanoparticles. 4) Detection of labelled RPA product immobilized on the lateral flow strip is performed by visual inspection using the naked eye. Total reaction time is approximately 35 minutes. Figure made using *BioRender*.

The RPA assay was performed as per manufacturer instructions with slight modifications: each 50µL RPA reactioncontained 29.5 μl RPA-nfo rehydration buffer, 7.2 μl water, 2.5 μl 280 nM MgOAc, 2.1 μl forward primer (10 μM) and 2.1 μl reverse primer (10 μM), 0.6 μl of probe (10 μM), 1 μl (200ng/μl) human DNA (for non-clinical samples) and 5 μl DNA template (or proportional equivalent). Reactions were incubated at 39°C for 30 min with brief intermittent vortexing after four minutes. After amplification, 2 μL of each reaction was immediately diluted with 98 μL of wash buffer in an Eppendorf tube, and a disposable lateral flow strip (Ustar Biotechnologies Ltd, Hangzhou, China REF: D001-03) was dipped into the tube. The result was recorded after 5 minutes, although results are usually apparent earlier. A test was considered positive when both the test line and control line visualized by the naked eye; and negative only when the control line is visible. When no line appeared, the result was considered invalid. Samples with invalid results were retested.

### Assay optimization

Both TwistDX basic and TwistDX nfo kits were used during optimization of reaction conditions before evaluation of assay performance. TwistDX basic products were denatured using heat (60°C for 10 minutes) or purified using commercially available DNA purification kit (Qiagen, Germantown, USA) and visualized by 3% agarose gel electrophoresis. The RPA reaction was performed according to manufacturer’s protocol except with temperature ranging from 34-40 °C for 30 min. For non-clinical samples, 18S rRNA plasmid DNA was combined with 200ng of human DNA to simulate human sample collection.

### Assessment of assay performance

To determine analytical sensitivity and specificity of the assay, 18S rRNA plasmid DNA of *P. malariae* (MRA-179, Lot:70031271), *P. falciparum* (MRA-152-G, Lot:59201399), *P. vivax* (MRA-178,Lot:58067149), and *P. ovale* (MRA-180,Lot:70043212) obtained from MR4 (BEI Resources, Manassas, USA) was used. To determine limit of detection, Pm 18S rRNA plasmid DNA was diluted from 10^6^ to 10^−1^ copies/μL. Analytical specificity was evaluated using 18S rRNA DNA from other *Plasmodium* species at high concentration (Pf and Pv at 10^6^ copies/µL, Po at 10^3^ copies/µL). Each reaction was performed with a minimum of four replicates.

Clinical sensitivity and specificity were evaluated using genomic DNA from patients infected with Pm, Pf, Po, and/or Pv collected as part of studies in Tanzania, Democratic Republic of Congo (DRC), and Ethiopia [8-10]. For all studies, participants provided informed consent or parental consent was obtained at the time of enrollment. DNA was extracted from dried blood spot samples using Chelex-100, and evaluated for Pm, Pf, Po, and Pv infection using real-time PCR targeting the 18S rRNA gene as previously described [8].

## RESULTS

### Selection of primers, probe, and optimized conditions

Fifteen primer pairs were evaluated for the current Pm RPA assay (listed in **Supplementary Table 1**). The length of the primers tested ranged between 30 and 41 nt, with melting temperature ranging from 51 to 62.4 °C and GC content ranging from 20 to 41%. The best performing primers and probe during initial testing using TwistDX basic kits were selected for further evaluation and validation using the TwistDx nfo kit: primers AJMP_7: 5’-ATAACATAGTTGTACGTTAAGAATAACCGC-3’ (forward), AJMP_30: 5’- ATATATAATACTTCGATTAGTTGAGTACCT-3’ (reverse), and probe AJMP_42: 5’- GTTGTACGTTAAGAATAACCGCCAAGGCTTTATTTTTTCTGTTAC-3’. Primer and probe modifications are depicted in **Supplementary Table 2**. The optimized RPA assay reaction consisted of two-steps: 1) RPA assay in a heat block at 39 °C for 30 min, and 2) lateral flow assay detection with visualization of results after 5 minutes (35-minute total reaction time, not including DNA extraction).

### Analytical sensitivity and specificity

The assay’s analytical sensitivity approached the sensitivity of Pm-specific PCR assays [8] and achieved perfect analytical specificity (**Figure 2A-2B**). When tested against serial dilutions of Pm 18S rRNA plasmid DNA, the lower limit of detection was between 10 and 100 copies/µL (∼1.7-17 genome equivalents/µL, assuming six copies of 18S rRNA per genome [11]), with all replicates containing ≥10 Pm copies/µL detected except for a single replicate. Specificity assessed using high-concentration plasmid DNA for Pf (10^6^ genomes/µL), Pv (10^6^), and Po (10^3^) confirmed 100% analytical specificity.

**Figure 2.**
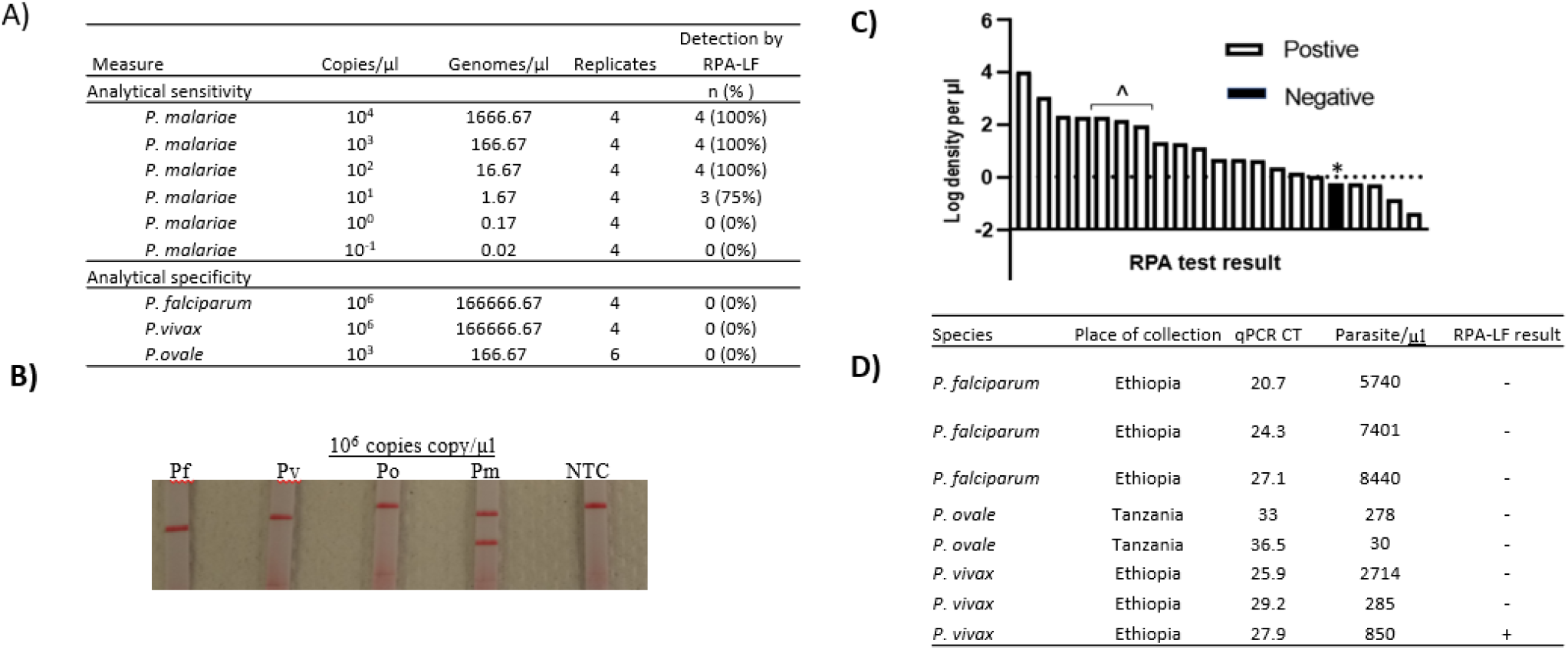
Sensitivity and specificity of the Pm RPA-lateral flow (RPA-LF) assay. A) Analytical sensitivity and specificity determined using serially diluted 18S rRNA plasmid copies/µL (equivalent to approximately 1,700-0.17 genome equivalents/µL). B) Example lateral flow read-out by species (Pf, *P. falciparum;* Pv, *P. vivax*; Po, *P. ovale* spp.; Pm, *P. malariae*; NTC, no-template control). C) Clinical sensitivity determined using 21 Tanzania field samples with qPCR-confirmed *P. malariae* infection with a range of qPCR-determined parasite densities (log_10_). One low-density case was missed (*). Three cases of Pm+Pf co-infection were included (^). D) Clinical specificity determined using Tanzania and Ethiopia field samples with qPCR-confirmed Pf, Po, and Pv.

### Clinical sensitivity and specificity

Clinical sensitivity and specificity assessed using field samples from Tanzania and Ethiopia were excellent, with only a single false-negative and false-positive, respectively (**Figure 2C-2D**). The assay’s clinical sensitivity compared to qPCR was 95.2%, detecting 20 of 21 Pm qPCR-confirmed field samples (17 mono-infected and three mixed Pf and Pm co-infection) from Tanzania with a range of parasite densities down to 10^0^ and 10^−1^ parasites/µL, corresponding to Ct values ranging 22-41. We observed a false-negative result in a single Pm field sample with parasite density of 0.5 parasites/µL, below the assay’s analytical limits of detection. Clinical specificity was 88% during testing against three Pf, two Po, and three Pv field samples from Tanzania and Ethiopia. One false-positive result was observed in a Pv field sample from Ethiopia, with a faint positive band that was not visualized during repeat testing.

## DISCUSSION

We describe a new Pm RPA-lateral flow assay with strong performance when applied to laboratory and field samples, and potential for future field use. Species-specific point-of-care assays for neglected *Plasmodium* species like *P. malariae* are not currently available. PCR assays for these species require sophisticated equipment and materials that are not available in many settings where malaria is endemic. Our Pm RPA-LF assay provides a simple, sensitive, and specific option to detect Pm in laboratories in low-resource settings.

The Pm-specific RPA-LF assay achieved limits of detection 10-100 fold better than existing pan-*Plasmodium* lactate dehydrogenase rapid diagnostic tests, with a lower limit of detection of 1.7-16.7 versus 100-1,000 parasites/µL, respectively[12]. The assay had perfect analytical specificity and yielded only a single false-negative during testing against field samples. The false-negative result was observed in a sample with low parasite density below the assay’s limit of detection. While the assay detected several other field samples with lower parasite densities, its performance against low-density samples ≤10 parasites/µL is expected to be less robust than higher-density samples, but this is also the case with qPCR. The assay had excellent specificity, achieving perfect results versus high-concentration plasmid DNA. Its single false-positive during testing of field samples involved a faintly positive test band that was not observed during repeat testing as part of post hoc analysis. This suggests that the initial false-positive call was likely a result of contamination during sample processing and RPA-lateral flow testing. RPA provides a rapid method of amplification, reportedly faster than other isothermal amplification methods [13], that can be performed at/near human physiological temperatures. Our assay had a total reaction time of less than 40 minutes and performed best at 39°C, suggesting opportunities for future use without a heat block (ex: incubation in the axilla or near a simple stove), though we did not test these approaches here.

Several limitations must be overcome before our assay is ready for point-of-care use. First, reagent stock outs hampered RPA-lateral flow diagnostic development throughout the COVID-19 pandemic and have not been rectified. Alternatives to TwistDx’s proprietary approaches and kits are now becoming available [14]. Second, our assay requires DNA input and cannot be applied directly to whole-blood samples in its current form. DNA extraction is required prior to performance of the RPA-lateral flow assay and adds time and complexity to sample processing. Advances in simple DNA extraction methods promise rapid turnaround time (<10 minutes), but nonetheless add complexity to the workflow. One-pot reactions that eliminate this requirement would simplify assay requirements and improve assay performance. Third, though our assay outperforms pan-*Plasmodium* RDTs and was on par with conventional real-time PCR, it could still miss low-density Pm infections. This is important because Pm infections have lower parasite densities than Pf and can be missed by commonly used PCR assays. However, it is still likely to pick up the majority of Pm parasitemias detectable by qPCR as well as those contributing to febrile illness [15]. Thus, our assay fills a key gap in the malaria diagnostic portfolio and outperforms pan-*Plasmodium* RDTs.

Evidence is limited about *P. malariae* – including its true prevalence and distribution and the extent of its propensity to cause chronic carriage and clinical disease. Simpler diagnostic tools that are closer to the point of care could help resolve some of these questions. They may also contribute to efforts to achieve malaria elimination, as all malaria species need to be detected and addressed. Our Pm-specific RPA-LF assay provides a useful tool for countries seeking to address a key neglected malaria species known for chronic parasitism in low resource settings.

## Data Availability

All data produced in the present study are available upon reasonable request to the authors

## Author contributions

JBP, JTL, and AA. conceptualized the study. AA, BN, AK, FP, and JBP data collection, fieldwork, and supervision. AA, KKW, CMH, CHC, and MM laboratory analyses. AA wrote the first draft. All authors reviewed and approved the final draft.

## Research Ethics

The study was ethically exempted by the Office of Human Research Ethics, University of North Carolina, Chapel Hill (Ref. 278853).

## Acknowledgements

The authors thank the study teams involved in sample collection and participants in the parent field studies.

