## Supplementary material for "Detection of *P. malariae* using a new rapid isothermal amplification lateral flow assay": Suplementary table 1 and 2

**Supplementary Table 1**. List of RPA Primers investigated for P. malariae detection in the study

| Name | sequence | Type | Length | Tm (^0^C) | GC (%) | Gene | References |
| --- | --- | --- | --- | --- | --- | --- | --- |
| AJM_01 | TATATGAGTGTTTCTTTTAGATAGCTTCCTT | Left (F) | 31 | 59.9 | 29 | 18s | Designed /modified |
| AJM_02 | TATGCCGACTAGGTGTTGGATGATAGAGTAAA | Left (F) | 32 | 67.9 | 40.6 | 18s | Designed /modified |
| AJM_03 | AATCCTACTCTTGTCTTAAACTAGTGAGTTTCC | Right (R) | 33 | 63 | 36.4 | 18s | Designed /modified |
| AJM_04 | ATATATGAGTGTTTCTTTTAGATAGCTTCCTTC | Left (F) | 33 | 61 | 30.3 | 18s | Designed /modified |
| AJM_05 | GTTTCTTTTAGATAGCTTCCTTCAGTACCTTAT | Left (F) | 33 | 62.2 | 33.3 | 18s | Designed /modified |
| AJM_06 | TTCTTTTAGATAGCTTCCTTCAGTACCTTAT | Left (F) | 32 | 61.1 | 31.2 | 18s | Designed /modified |
| AJM_07 | ATAACATAGTTGTACGTTAAGAATAACCGC | Left (F) | 30 | 56 | 33 | SSUrRNA_Pm_gene (M54897.1) | Snounou et al., 1993 |
| AJM_08 | AAAATTCCCATGCATAAAAAAT TATACAAA | Right(F) | 30 | 51 | 20 | SSUrRNA_Pm_gene (M54897.1) | Designed /modified |
| AJM_09 | AGTAATGCTTTGTATATTTATAACATAGTTG | Left (F) | 31 | 55.8 | 22.6 | SSUrRNA_Pm_gene (M54897.1) | Designed /modified |
| AJM_10 | AACACTCTAATTTACTCAAAGTAACAAAATTC | Right(R) | 32 | 59.5 | 25 | SSUrRNA_Pm_gene (M54897.1) | Designed /modified |
| AJM_11 | CTTATATATGAGTGTTTCTTTTAGATAGCTTCC | Left (F) | 33 | 60.1 | 30.3 | SSUrRNA_Pm_gene (M54897.1) | Designed /modified |
| AJM_12 | CTATTAATCTGTCAATCCTACTCTTGTCTTAAA | Right(R) | 33 | 61.1 | 30.3 | SSUrRNA_Pm_gene (M54897.1) | Rutledge et al., 2017 |
| AJM_13 | CTTATATATGAGTGTTTCTTTTAGATAGCTTCC | Left (F) | 33 | 60.1 | 30.3 | SSUrRNA_Pm_gene (M54897.1) | Goman et al., 1991 |
| AJM_14 | ATTAATCTGTCAATCCTACTCTTGTCTTAAACT | Right(R) | 33 | 61.6 | 30.3 | SSUrRNA_Pm_gene (M54897.1) | Designed /modified |
| AJM_15 | ATATATGAGTGTTTCTTTTAGATAGCTTCCTTC | Left (F) | 33 | 61.3 | 30.3 | SSUrRNA_Pm_gene (M54897.1) | Designed /modified |
| AJM_16 | CTATTAATCTGTCAATCCTACTCTTGTCTTAAA | Right(R) | 33 | 61.1 | 30.3 | SSUrRNA_Pm_gene (M54897.1) | Designed /modified |
| AJM_17 | CTTATATATGAGTGTTTCTTTTAGATAGCTTCC | Left (F) | 33 | 60.1 | 30.3 | SSUrRNA_Pm_gene (M54897.1) | Designed /modified |
| AJM_18 | AGCTATTAATCTGTCAATCCTACTCTTGTCTTA | Right(R) | 33 | 62.4 | 33.3 | SSUrRNA_Pm_gene (M54897.1) | Designed /modified |
| AJM_19 | CTTATATATGAGTGTTTCTTTTAGATAGCTTCC | Left (F) | 33 | 60.1 | 30.3 | SSUrRNA_Pm_gene (M54897.1) | Designed /modified |
| AJM_20 | TTAATCTGTCAATCCTACTCTTGTCTTAAACT | Right(R) | 32 | 61.5 | 31.2 | SSUrRNA_Pm_gene (M54897.1) | Designed /modified |
| AJM_21 | ACATTCTTATATATGAGTGTTTCTTTTAGATAGCTTC | Left (F) | 37 | 58 | 27 | SSUrRNA_Pm_gene (M54897.1) | Designed /modified |
| AJM_22 | GTTACAAATAATTATAAAACTTAACACGTACT | Left (F) | 32 | 56.2 | 21.9 | PmUG01_13030700 | Designed /modified |
| AJM_23 | GTTTAAATCTCTACTTATGCTTAATATATTCTC | Right(R) | 33 | 56.5 | 24.2 | PmUG01_13030701 | Designed /modified |
| AJM_24 | ACTGTTACAAATAATTATAAAACTTAACAC | Left (F) | 30 | 54.1 | 20 | PmUG01_13030702 | Designed /modified |
| AJM_25 | TTTAAATCTCTACTTATGCTTAATATATTCTC | Right(R) | 32 | 55.9 | 21.9 | PmUG01_13030703 | Designed /modified |
| AJM_26 | ACTGTTACAAATAATTATAAAACTTAACAC | Left (F) | 30 | 54.1 | 20 | PmUG01_13030704 | Designed /modified |
| AJM_27 | TTAAATCTCTACTTATGCTTAATATATTCTC | Right(R) | 31 | 54.8 | 22.6 | PmUG01_13030700 | Designed /modified |
| AJM_28 | AATATATAATACTTCGATTAGTTGAGTACCT | Left (F) | 31 | 55.8 | 25.8 | PmUG01_13030700 | Designed /modified |
| AJM_29 | TACTATTTAGAAGAACATGAATAAGAATTT | Right(R) | 30 | 55.5 | 20 | PmUG01_13030700 | Designed /modified |
| AJM_30 | ATATATAATACTTCGATTAGTTGAGTACCT | Right(R) | 30 | 53 | 27 | PmUG01_13030700 | Designed /modified |
| AJM_31 | TCAACACGGGGAAACTCACTAGTTTAAGA | Left (F) | 41 | 59 | 41 | SSUrRNA_Pm_gene (M54897.1) | Designed /modified |

**Supplementary Table 2**

Primers and probe set and modifications used in the assay: 1) forward primer an unlabeled 2) reverse primer biotinylated on the 5’ end and 3) 5’-FAM-labelled probe with an abasic residue and 3’ blocker modification.

| Name | Type | Modified sequence |
| --- | --- | --- |
| AJMP_7 | Forward primer | 5’-ATAACATAGTTGTACGTTAAGAATAACCGC-3’ |
| AJMP_30 | Reverse primer | 5’-/5Biosg / ATATATAATACTTCGATTAGTTGAGTACCT-3’ |
| AJMP_42 | Probe | 5’-/FAM-dT/GTTGTACGTTAAGAATAACCGCCAAGGCTT/idSp/TATTTTTTCTGTTAC/3SpC3/-3’ |
